# Sleeping Through Menopause: study protocol of a randomized controlled trial of menopausal hormone therapy and online guided cognitive behavioural and circadian therapy for insomnia in perimenopausal women suffering from insomnia

**DOI:** 10.64898/2026.07.22.26358657

**Authors:** Francesca van Baarzel, Birit F.P. Broekman, Dorenda K.E. van Dijken, MenoPause Consortium, Eus J.W. van Someren, Anne-Sophie C.A.M. Koning

**Affiliations:** Amsterdam University Medical Center, Department of Psychiatry, Vrije Universiteit van Amsterdam, Amsterdam, The Netherlands; OLVG Hospital, Department of Psychiatry and Medical Psychology, Amsterdam, The Netherlands; OLVG Hospital, Department of Gynaecology, Amsterdam, The Netherlands; Jan van Goyenkliniek, Amsterdam, The Netherlands; VU University Amsterdam, Department of Integrative Neurophysiology, Center for Neurogenomics and Cognitive Research (CNCR), Amsterdam, The Netherlands; Netherlands Institute for Neuroscience, Department of Sleep and Cognition, Amsterdam, The Netherlands

**Author notes:** Corresponding author: Francesca van Baarzel. A list of authors and their affiliations appears at the end of the paper.

**Keywords:** Insomnia, Perimenopause, Menopausal hormone therapy, Cognitive behavioural circadian therapy, Cognitive behavioural therapy for insomnia, Circadian rhythm support

## Abstract

**Background:** About half of the women experience insomnia during the perimenopause, a transitional phase characterised by fluctuations in sex hormones. Insomnia is known to increase the risk of other mental health problems. Therefore, addressing insomnia could potentially prevent or alleviate mental health issues. Unfortunately, it is currently unknown which treatment is most effective in alleviating insomnia during the perimenopause. The Sleeping Through Menopause trial is among the first study to investigate side-by-side two interventions; menopausal hormone therapy (MHT) and combined cognitive behavioural and circadian therapy for insomnia (CBCTi). The primary aim of this study is to evaluate the effectiveness of CBCTi and MHT on insomnia symptoms.

**Methods:** We plan a repeated measurement, randomised controlled trial (RCT) in perimenopausal women who experience insomnia and climacteric symptoms. The study uses a web-based, nationwide recruitment approach, allowing participants from across the Netherlands to self-enroll via an open-access research website. After informed consent, 222 women will be randomly allocated to one of four conditions in a two-by-two treatment design: 1. MHT, 2. CBCTi, 3. MHT + CBCTi and 4. control. MHT consists of transdermal estradiol patches and progesterone tablets in standard doses. CBCTi addresses sleep and circadian behaviour and cognitive restructuring. The primary study outcome is the change in insomnia symptoms assessed with the Insomnia Severity Index (ISI) after 8 weeks (T1) and 15 weeks (T2) relative to baseline (T0). Secondary outcomes are other subjective sleep quality indicators, objective sleep estimated from actigraphy and headband EEG, changes in climacteric symptoms, hot flash frequency estimated from wrist electrodermal activity, other mental health complaints, and wellbeing and daytime functioning. Furthermore, baseline personal characteristics will be assessed, including the history of hormone-related mental health problems.

**Discussion:** This study addresses the gap in research regarding optimal treatment for insomnia during the perimenopause, and how treatments alleviate or prevent other mental health problems and climacteric symptoms including hot flashes.

**Trial Registration:** The study was prospectively registered in the National Library of Medicine on May 13^th^ 2024 as ‘Improving Sleep and Mood in Peri-Menopause’ (NCT06306404, https://clinicaltrials.gov/study/NCT06306404?cond=NCT06306404&rank=1).

## BACKGROUND

The menopause is an important health transition in female aging and is defined by the last menstrual cycle in women’s life. The 5-8 years period preceding menopause and one year after, the perimenopause, is characterised by fluctuations in sex hormones. This leads to lasting irregular menstrual cycles, heavier or lighter bleeding and skipped periods (1). Many women (approximately 60% to 80%) experience physical and mental symptoms during the perimenopause, with around 30% reporting them as severe or significantly impacting their quality of life (2–4). In particular, hot flashes are the most frequently reported climacteric symptom, affecting up to 68% of women (5). These sudden sensations of heat, often accompanied by sweating and a rapid heartbeat, can disrupt sleep and daily functioning. Furthermore, the perimenopause forms a vulnerable period for the development of mental health disorders. Perimenopausal women have a 2-5 times increased risk for new onset and recurrent depression (6–9) and about 50% experience insomnia symptoms, particularly night-time awakenings (10).

Insomnia is increasingly recognized as a contributing factor to the development of mental disorders rather than simply as a symptom of these conditions (11, 12). Recent research on the role of sleep in overnight emotion regulation suggests that poor sleep quality may disrupt emotional processing, potentially leading to emotional distress and the development of mental disorders (10, 11, 13–16). Although insomnia is a disorder that can occur independently, it is often comorbid with other mental disorders, such as depression and anxiety. Poor sleep quality could be an important maintaining factor in mental disorders as insomnia potentially aggravates mental disease state, worsens prognosis, impedes treatment response, and promotes relapse after recovery of mental disorders (17–21).

Despite that most studies find that sleep problems are one of the most common complaints reported by perimenopausal women to their healthcare providers (5, 22), specific insomnia treatment is not considered an integral part of menopausal care. In fact, it is not typically included in general mental health care as well. This is a missed opportunity since cognitive behavioural therapy for insomnia (CBTi) is an effective sleep intervention. CBTi addresses lifestyle factors of sleep, physical activity, stress, smoking, use of alcohol and circadian rhythm regularity (23–25). Of note, when applied in people with major depressive disorder (MDD), CBTi not only ameliorated insomnia itself, but also depressive symptoms (26–30) and reduced the risk of future depressive episodes (30, 31). Especially CBTi in combination with circadian therapy (CT), also called circadian rhythm support (CRS) has been shown to prevent worsening of depressive symptoms (32). CT addresses the regularity of the circadian rhythm by controlling light exposure, temperature and movement. The integration of CBTi and CT will be referred to as cognitive behavioural circadian therapy (CBCTi) in this study.

Next to CBCTi, some studies show that menopausal hormone therapy (MHT) may also relieve insomnia symptoms by reducing nocturnal hot flashes, or night sweats, which in themselves cause sleep disturbances (33). MHT consists of estradiol in combination with progesterone or progestin (except for women with a hysterectomy and no history of endometriosis). Preclinical and clinical data support the benefits of estrogen-based therapies in improving both depressive symptoms and sleep quality (34, 35). Additionally, allopregnanolone, a metabolite of progesterone, is believed to decrease vigilance by exerting effects on GABA receptors in the brain, resulting in a sedative effect that facilitates earlier sleep onset (36).

It is currently unclear whether MHT is more effective in decreasing insomnia symptoms during perimenopause compared to the current first-line insomnia treatment, CBCTi. Women who experience bothersome nightly hot flashes or who are historically sensitive to hormonal fluctuations, as evidenced by conditions such as Premenstrual Syndrome (PMS) and Premenstrual Dysphoric Disorder (PMDD), may particularly benefit from MHT. Whereas women whose insomnia is primarily driven by behavioural and psychological factors, might benefit mostly from CBCTi. The combination of both therapies is expected to be the most effective, as it reduces nightly awakenings and tackles any other sleep obstructing behaviour or thought patterns.

The first aim of this study is to evaluate the effectiveness of MHT and guided online CBCTi in reducing insomnia severity in women suffering from both insomnia and climacteric symptoms, with the Insomnia Severity Index (ISI) as primary sleep outcome measure. Secondly, the effect of both therapies will be evaluated on other sleep outcomes, climacteric symptoms including hot flashes, and various mental health outcomes, like depressive symptoms and anxiety. The third aim is to explore treatment effect modification based on individual characteristics, including a history of hormone-related mental health issues like PMS and PMDD.

## METHODS ANDS ANALYSIS

### Research design

This web-based multi-centre repeated measures randomized controlled trial (RCT) will be performed in collaboration with Amsterdam University Medical Center, the Netherlands Institute for Neuroscience, and OLVG hospital. This study aims to investigate the effectiveness of MHT and CBCTi in perimenopausal women with insomnia and climacteric symptoms. This trial was approved by the Medical Ethics Committee of the Amsterdam University Medical Center (NL87156.018.24), and the board of directors of all participating centres. The study was prospectively registered in the National Library of Medicine on May 13^th^ 2024 (NCT06306404). All participants will provide informed consent. The SPIRIT Standard Protocol Items: Recommendations for Interventional Trials is provided as part of the supplementary information (S1).

### Participants

This study uses a web-based, nationwide recruitment design, allowing participants from across the Netherlands to self-enroll via an open-access research website, the Netherlands Sleep Registry (NSR) (http://www.slaapregister-menopauze.nl). Participants on waiting lists for specialised menopause clinics at OLVG hospital, Amsterdam, The Netherlands, will be approached by a healthcare professional to consider self-enrolment via NSR. Women are eligible to participate in the study if they meet the following criteria: (1) they are between 40 and 55 years old, (2) have insomnia severity index (ISI) of 10 or higher, (3) have green climacteric scale (GCS) score of 13 or higher, (4) have self-considered capability of completing online questionnaires and diaries in Dutch or English and (5) their last menstruation is less than 12 months ago. Women are excluded from the study if they (1) have undergone CBTi in the past year, (2) already use MHT, (3) diagnosed with bipolar or psychotic disorders, (4) have contra-indication for the use of MHT (e.g. breast cancer, meningioma, liver diseases, trombo-embolic diseases), (5) have undergone a hysterectomy and/or oophorectomy (6) use medication that interact with MHT (e.g., lamotrigine, aromatase), (7) use hormonal contraceptives, (8) have an alcohol or drug dependency (scores of ≥ 20 on the AUDIT and ≥25 on the DUDIT), (9) use drugs known to interfere with cytochrome P450 enzyme (CYP) 3A4 and (10) are known with hypersensitivity to the excipients in the estradiol patch and progesterone capsule.

### Randomization and blinding

Participants will be allocated to one of four groups (1. CBCTi, 2. MHT, 3. CBCTi + MHT, 4. Control group). An independent researcher will create an allocation scheme using a computerized random number generator. Simple randomization will be applied for the first 10 participants. In order to ascertain balanced groups, subsequent participants will be assigned according to covariate adaptive randomization (37, 38) scripted in R (R Core Team, 2024) with covariates age, time of year of inclusion, baseline severity of insomnia, baseline severity of depressive symptoms and baseline severity of climacteric symptoms. Due to the nature of the interventions, the participants and study team members will not be blinded to the condition a participant is allocated to.

### Study outline

Women interested in participating will complete an online screening questionnaire through the web-based NSR platform. After filling out the questionnaire, eligible women will be contacted by the researchers to re-check the in- and exclusion criteria and to provide extra study information. After a consideration period, participants can indicate whether they want to participate. Participants will visit the research clinic based in OLVG hospital Amsterdam, where they sign informed consent after which they will be allocated to one of four conditions.

Participant characteristics will be assessed in the online baseline questionnaire. Repeated assessments will take place at week 1 (T0), at week 8 (T1) and at week 15 (T2). An overview of the measurements at each time point is shown in Table 1.

**Table 1.**
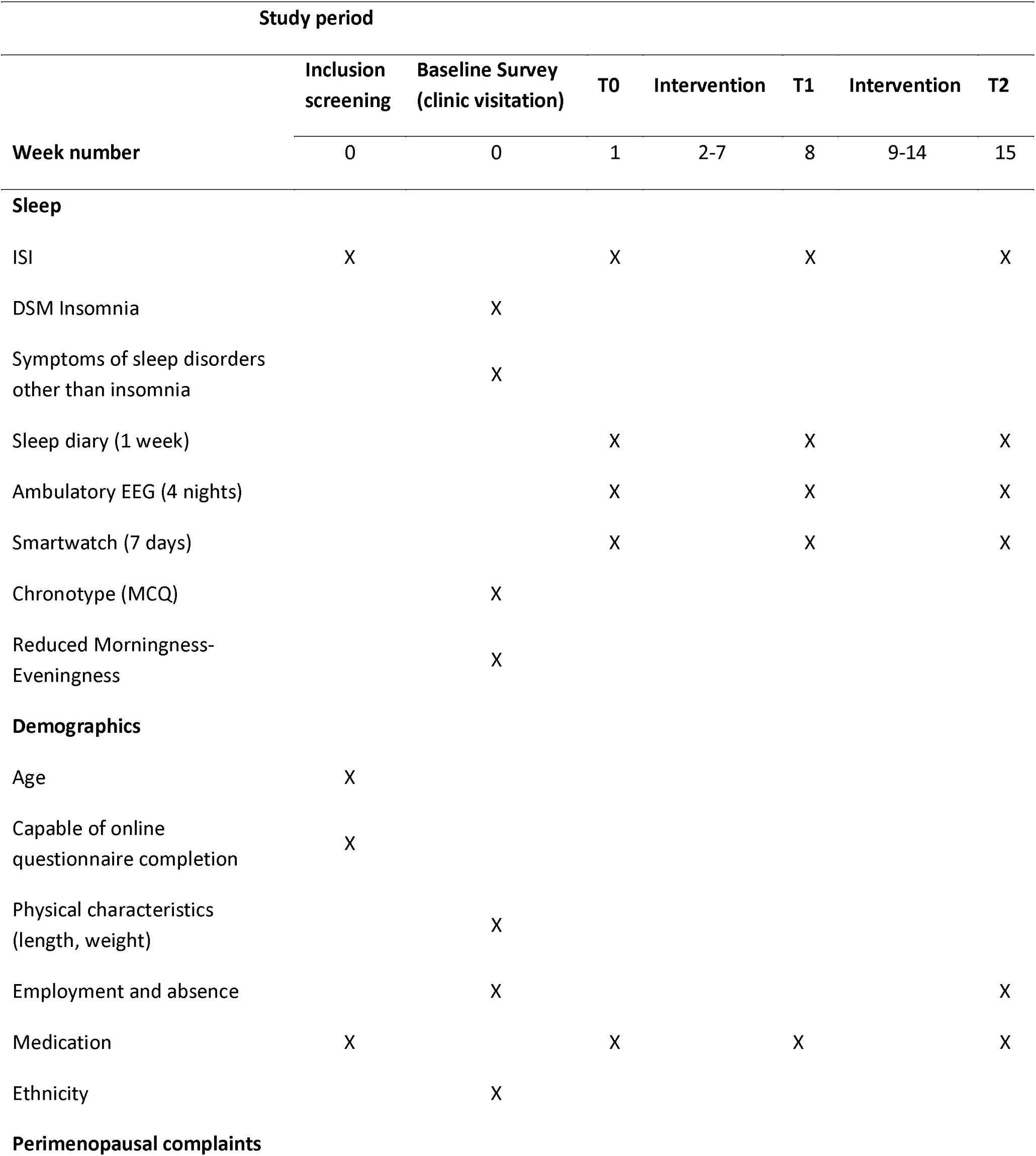
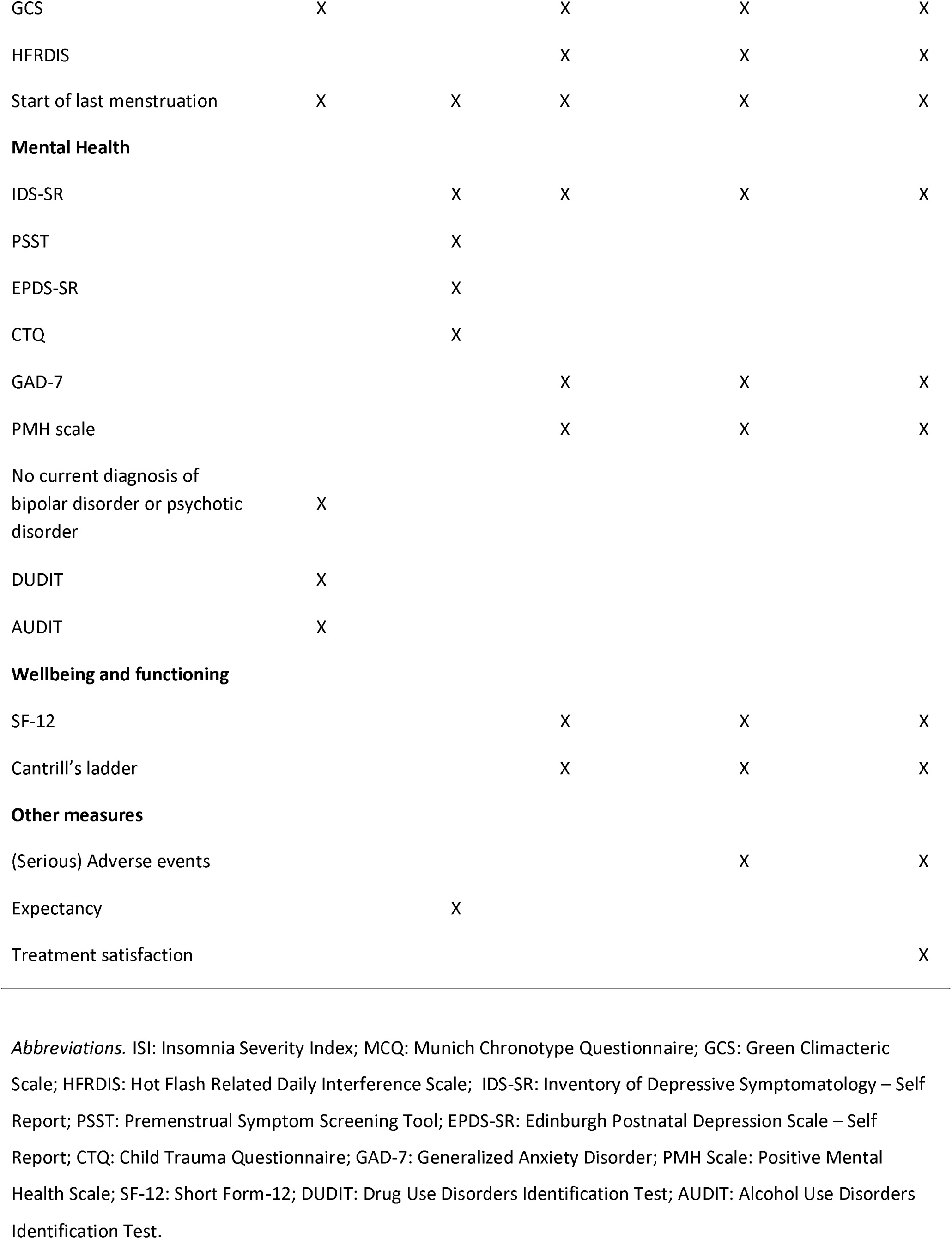
Study Schedule, intervention and assessments.

In addition, ambulatory Z-max triple electrode EEG headbands (Hypnodyne) will be used to assess brain activity during sleep, allowing for the assessment of sleep stages and sleep architecture, at home for four nights during each timepoint. Participants will also be given a smartwatch (Embraceplus) that can measure among others, heartrate, electrodermal activity, and physical activity for seven days at each timepoint. Participants are free to opt out of EEG and smartwatch measurements at any time without consequences for study participation, treatment or otherwise.

### Intervention

#### Cognitive Behavioural Circadian Therapy for insomnia (CBCTi)

The web-based guided sleep intervention consists of 5 modules, in which cognitive behavioural therapy for insomnia (CBTi) and circadian therapy (CT) are incorporated. CBTi entails sleep hygiene, lifestyle factors, sleep restriction, stimulus control, cognitive restructuring and relaxation. CT provides supporting tools to strengthen the regularity of the circadian rhythm, e.g, increase daylight exposure. For an overview of the 5 modules see Table 2. The online CBCTi is an adaptation from the treatment originally developed for primary care by Vrije Universiteit Amsterdam and is currently available in daily clinical practice for treating insomnia and regarded as first line treatment (24, 25, 39, 40).

**Table 2.** Overview of the CBCTi across five modules.

| Session | Theme | Procedures |
| --- | --- | --- |
| 1 | Sleep education and implementation of strategies | General guidelines about health and environmental factors influencing sleep, |
|  |  | including physical activity (CT), are provided and how to implement newly learned strategies according to intention to implement principle (CBTi). |
| 2 | Light exposure and sleep restriction | Adjusting light exposure to positively influence sleep (CT) and introduction of sleep restriction to retrain the sleep pattern (CBTi). |
| 3 | Relaxation and regulating body temperature | Incorporating relaxation techniques in everyday life (CBTi) and providing guidelines in regulating body temperature to promote sleep (CT). |
| 4 | Cognitive restructuring (i.e. changing misconceptions about sleep) | Identifying sleep obstructing thought patterns and restructuring them (CBTi). |
| 5 | Future plans and relapse prevention | Dealing with setbacks by introducing techniques from acceptance commitment therapy (CBTi). |

Alongside the CBCTi modules, a consensus sleep diary is daily kept which is a validated method for sleep self-monitoring (41). Adhering to the intervention takes an estimated 10 to 30 minutes a day. Participants are asked to complete one module per week, with a one-week extension allowed, totalling six weeks for the intervention. CBCTi will be delivered through the NSR, which can be accessed from PC, smartphone and tablet. Participants can log in with their email address and password to access the CBCTi modules, digital sleep diary, questionnaires and a secure communication channel for both participants and their assigned therapists. Participants receive text-based feedback from a CBCTi trained therapist after the completion of each module. Participants can opt for a phone/video call with their therapist if they need extra guidance. All trained therapists receive monthly supervision from a clinical psychologist and intervision between therapists.

### Menopausal hormone therapy

MHT is a standard medical treatment that involves the use of hormones to reduce climacteric symptoms, in particular vasomotor symptoms (VMS), occurring during perimenopause and to reduce the risk of certain long-term health conditions associated with menopause such as osteoporosis and cardiovascular health problems. MHT will be administered according to the updated NICE guidelines (42). It consists of estradiol patches (Systen, 50mcg) and progesterone tablet (Utrogestan, 200mg). The estradiol patches will be applied to the skin twice per week over the course of 15 weeks. The progesterone tablets will be taken orally once daily for two weeks, to protect the uterus endometrium from the proliferative (thickening) effects of estrogen, followed by a two-week break.

### Concomitant care and withdrawal

Participants are not allowed to receive CBCTi or MHT elsewhere during the trial period. Participants may withdraw from the study at any time, without providing a reason. All data collected up till the withdrawal will be used for analysis. After completion of the trial, participants will be notified of the results.

### Outcome measures Primary outcome measure

The primary outcome is the difference between the groups in the within-subject change in insomnia severity at T1 (8 weeks) and T2 (15 weeks) relative to T0 (week 1). Insomnia severity will be assessed with the Insomnia Severity Index (ISI) on these three timepoints. The ISI consists of 7 items, scored on a 5-point Likert scale (0= not at all, 4= extremely), with scores ranging from 0-28 (higher scores indicating a higher insomnia severity) (43). The ISI has adequate internal consistency, is sensitive to changes and has been validated for online use (40, 43, 44). The clinical relevance of the decrease in ISI scores will be explained in two ways: 1) reporting the reliable change index (45), 2) discussing the percentage of participants for which the ISI scores remain stable or decreased according to earlier defined clinical relevance cut-off scores of -8.4 points (40) and -6 points (46).

### Secondary outcome measures Sleep outcomes

Secondary subjective sleep outcomes will be calculated from the relevant items of the consensus sleep diary (CSD) for seven nights in each timepoint (T0, T1, T2) (41). This includes sleep efficiency, sleep onset latency, wake after sleep onset, number of awakenings, terminal wakefulness, total sleep time, and subjective sleep quality. Objective secondary sleep outcomes, which are collected using the ambulatory EEG headband for four consecutive weeknights in each timepoint (T0, T1, T2), include sleep continuity- and sleep architecture measures.

### Climacteric symptoms

The Greene Climacteric Scale (GCS) will be used to assess differentiation in symptoms and severity of climacteric symptoms (47). The GCS is a self-administered tool and consists of 21 symptoms, each symptom is rated on a 4-point Likert scale (0=mild, 3=severe). The symptoms cover four domains: (1) psychological domain (items 1–11), divided into anxiety (items 1–6) and depression (items 7–11), (2) somatic symptoms (items 12–18), (3) vasomotor symptoms (items 19 and 20), and (4) sexual dysfunction (item 21). In addition, the Hot Flash Related Daily Interference Scale (HFRDIS) is used to assess the impact of hot flashes on daily life (48). The objective occurrence of nocturnal hot flashes will be estimated from a multisensory approach (skin conductance, skin temperature, heart rate and movement) assessed with the EmbracePlus smartwatch for seven days at each timepoint T0, T1 and T2 (49–51).

### Severity of depressive symptoms

To assess severity of depressive symptoms all participants fill out the well-validated Self Rated Inventory of Depressive Symptomatology (IDS-SR) at T0, T1 and T2. The IDS-SR is a 30 item questionnaire with a 4-point Likert scale (higher score indicates higher depression severity), with an excellent internal consistency (Cronbach’s alpha=0.94) (52). Sub-scores for atypical depression will be calculated based on characteristics of atypical depression according to the DSM-IV by calculating the sum of item scores for hypersomnia (item 4), mood reactivity (item 8, reverse scored to represent absence of anhedonia), increased appetite (item 12), weight gain (item 14), leaden paralysis (item 29) and interpersonal sensitivity (item 30) from the IDS-SR, resulting in a sum score ranging from 0 to 18 (53).

### Other mental health symptoms

Other mental health outcomes will be assessed with the Generalized Anxiety Disorder (GAD-7) questionnaire (54). This is a well validated and reliable questionnaire to measure core symptoms of generalized anxiety in the general population, including symptoms such as rumination and having trouble relaxing. We will also use the 9-item Positive Mental Health Scale (PMH-scale) (55). The PMH-scale has a high internal consistency, good retest-reliability and is sensitive to therapeutic change.

### Daytime functioning, lifestyle and wellbeing

Daytime functioning will be assessed with the Short Form-12 (SF-12v2) (56) This questionnaire assesses the quality of life on different domains, including physical functioning, role – physical, bodily pain, general health, vitality, social functioning, role – emotional and mental health. Wellbeing will be assessed by the 2-item Cantril’s Ladder of Life questionnaire (57).

### Individual differences in traits and life history

At baseline, demographics, baseline sleep, mental health and history of hormonal related physical and mental issues will be assessed. The Childhood Trauma Questionnaire (CTQ) is a 28-item retrospective self-report questionnaire and will be used to rate traumatic experiences during childhood (58). To indicate sensitivity for sex hormone fluctuations, the Premenstrual Screening Tool (PSST, 19 items) will be used (59). The lifetime Edinburgh Postnatal Depression scale (EPDS) is a validated questionnaire will be used to identify previous peri-partum depressions (60). The Munich Chronotype Questionnaire (MCTQ, 19 items) will be used to assess individual’s chronotypes (61). Lastly, the Reduced Morningness-Eveningness questionnaire (19 items) will be used to evaluate preference towards morning or evening (62).

### Sample size

(ANCOVA-type) Linear mixed effect regression models (LMMs) will be used to estimate treatment effects across T1 and T2, while controlling for individual differences in baseline values (T0). In a 2x2 design participants will either receive no intervention, CBCTi, MHT, or both treatments. Effects of CBCTi, MHT and their interaction will be simultaneously estimated in a single model. For each outcome, two repeated post-treatment assessments in 50 completing participants in each cell of the 2x2 design (total of n=200), yield a power of 0.80 at a significance of alpha=0.05 to detect small to medium-sized main effects of CBCTi and MHT of at least d=0.32 and to detect a medium-size interaction effect of at least d=0.64. Expecting a drop-out of 10%, the total number of participants to be recruited is n=222.

Effect modification will be evaluated in ancillary explorative analyses, that extend the basic overall analysis regression models with several regressors and their interactions with treatment conditions. The models estimate the extent to which the main and interaction effects of CBCTi and MHT on the primary and secondary outcome measures is modified by baseline severity of e.g. insomnia and hormonal-related physical and mental health issues in the past.

### Statistical analysis

The (ANCOVA type) LMM used to estimate treatment effects on the primary outcome measure of insomnia severity is shown below.

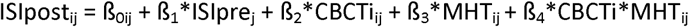

where ISIpost is the primary outcome, composed of insomnia severity post-treatment, assessed at two times (T1 and T2). ISIpre is the covariate of the individual’s insomnia severity pre-treatment, j refers to an individual (j = 1, .., n), i refers to the measurement within the individual (i=0, 1 for T1 and T2), β_0ij_ is the intercept; and MHT and CBCTi denote the dummy-coded assignment to the CBCTi and MHT conditions and their interaction. The coefficients β_2_, β_3_ and β_4_ estimate the fixed effects of CBCTi and MHT conditions and their interaction. The LMM model will be performed with the R package mmrm.

The same model as shown above applies to all other outcomes assessed at T0, T1 and T2, i.e. representing other sleep outcomes as well as the severity of symptoms depression, other mental health outcomes, climacteric symptoms and other sleep outcomes.

To explore effect modification by individual differences at baseline, interaction terms will be added to represent baseline severity of mental health and climacteric symptoms, sleep features, history of mental disorders and hormonal sensitivity and expectation of effectiveness as well as compliance during the intervention.

## DATA MANAGEMENT AND MONITORING

All data will be handled in accordance with the General Data Protection Regulation (GDPR). There are three main sites for storing data (1) electronic data on Amsterdam UMC endorsed data server, (2) hard-copy informed consent forms will be kept in a locked cabinet at the hospital site (OLVG) to ensure confidentiality and data, (3) questionnaire data at Netherlands Sleep registry. Participants will be pseudonymized, i.e. each participant will be identified by a study number. The key connecting names to the code will be safeguarded by authorised investigators. Results of the study will be kept for 25 years after publication and will then be destroyed. Coded data will be kept for a period of 25 years to be used for (re)analysis in (inter)national collaborations. If new questions arise or if our data can be used to answer new questions within the research field, other parties can request to use the data upon filling a Data Sharing proposal form. Monitoring is provided by the Clinical Monitoring Center (CMC) of Amsterdam UMC. The annual progress report will be combined with the annual safety report. Information will be provided on the numbers of subjects included and numbers of subjects that have completed the trial. The start date of the study will be reported separately to the Clinical Trial Information System (CTIS).

## SAFETY DSMB

A Data Safety Monitoring Board (DSMB) will not be assembled for this study. Both interventions are commonly used evidence-based therapies in the Netherlands.

### AEs and SAEs

We will monitor the occurrence of adverse events (AEs) using 3 inquiring questions in both conditions at timepoints T1 and T2. Patients will be asked about events such as traffic accidents and falls. We do not expect frequent adverse events based on prior research.

A serious adverse event (SAEs) that might be related to the investigational CBCTi treatment is the following: acute aggravation of psychiatric symptoms which necessitates additional intervention, such as a manic or psychotic episode. SAEs for MHT that are listed in the Summary of Product Characteristics (SmPC) for Utrogestan and Systen will not be reported. SAEs that will be monitored regarding MHT treatment are: (1) death; (2) life threatening events; (3) hospitalisation (but not elective admission); (4) persistent or significant disability or incapacity; (5) any other important medical event that did not result in any of the outcomes listed above. The investigator will report SAE to the sponsor without undue delay after obtaining knowledge of the events. The sponsor will provide an aggregated table of SAEs in the Annual Safety Report, which will be submitted in CTIS. We will monitor the occurrence of the abovementioned SAE throughout the trial period (T1 and T2).

## DISCUSSION

Sleeping Through Menopause is among the first study to date that investigates side-by-side CBCTi and MHT in perimenopausal women on improving insomnia symptoms. Addressing sleep problems and finding proper treatment for insomnia in perimenopausal women could have a large impact on their quality of life that extends further than solely improving sleep. Research suggests that when MHT is administered in the right way, at the right time, it could have long lasting beneficial effects on overall mental and physical health (63). Several other studies found that CBTi not only alleviate insomnia symptoms in menopausal women, but also improve depressive symptoms, overall well-being and other climacteric symptoms, such as hot flash interference (64–66). CBTi may thereby serve as a preventative intervention for severe climacteric symptoms and mood disorders and could also be considered a first-line treatment for women who are unwilling or unable to use MHT.

This study has several strengths. We aim to include a representative sample by employing lenient inclusion criteria and recruiting participants from the general population and via specialised menopause health clinics. This approach ensures a broad range of participants with varying levels of menopausal and insomnia symptoms, thereby enhancing the generalizability of the findings. The design of this study facilitates at home measurements of subjective and objective sleep parameters, as well as objectively measured hot flashes. This reduces the daily burden for the participants.

Additionally, the online guided CBCTi intervention might be preferred by participants reluctant to have face-to-face therapy.

The study also has some (potential) limitations. Firstly, this study is not sufficient powered for secondary analyses, rendering them exploratory in nature. Nevertheless, these results may still yield valuable insights and contribute to a deeper understanding of the data. Secondly, participants with severe symptomatology might not have the capacity to adhere to all the interventions and extra study requirements or prefer face-to-face therapy. Lastly, the MHT used in this study is only one of many ways MHT can be administered. There is a large global variety in MHT substrates (e.g., synthetic or bio-identic hormones), dose and way of administration (e.g., oral, gel, patches, spray). New products are developed rapidly as well, which cannot be included in this study.

## Data Availability

The data collected in this study will be available from the corresponding author upon reasonable request and after publication of findings.

## Abbreviations

CBCTi: Cognitive Behavioural Circadian Therapy
MHT: Menopausal Hormone Therapy
CBTi: Cognitive Behavioural Therapy for insomnia
CT: Circadian Therapy
CRS: Circadian Rhythm Support
ISI: Insomnia Severity Index
NSR: Netherlands Sleep Registry

## DECLARATIONS

### Ethics approval and consent to participate

The study has been approved by the Medical Research Ethics Committee of the Amsterdam University Medical Center (METC AUMC), and by the Institutional Review Boards of the participating sites. Perimenopausal women will only be included if they agree to participate and sign the informed consent. The study will be performed according to the principles of the Declaration of Helsinki (October 2013), Good Clinical Practice guidelines and in accordance with the Medical Research Involving Human Subjects Act (WMO). Any amendment to the protocol will be approved by the METC AUMC. Participant WMO insurance is provided by the Amsterdam UMC for all participating subjects and liability insurances are provided by all participating centres. Publication rights and ownership of data is secured in a clinical trial agreement with participating centers. Results of the study will be submitted for publication in a peer-reviewed journal.

### Consent for publication

Not applicable.

### Protocol versions

V8.0, September 5 2025: approved protocol

### Authors’ contributions

FvB: Writing – original draft; AK, BB, DvD, EvS: Writing – review & editing. All authors read and approved the final manuscript. MenoPause Consortium: applicants in the funding acquisition.

## Acknowledgements

Not applicable.

## MenoPause Consortium

Annegreet Vlug^4,7^, Annemieke Heijboer^8,9^, Astrid Bakker^10^, Birit F. P. Broekman^1,2^, Cindy L. Kelder^11,^ Dorenda K. E. van Dijken^3,4^, Eus J. W. van Someren^1,5,6^, Eveline Bruinstroop^12^, Fedde Scheele^13,14^, Fernando Rivadeneira^15^, Irene van Valkengoed^16^, Jacqueline Broerse^13^, Jeanine Roeters van Lennep^15,17^, John Dierx^18^, Karen Nieuwenhuijsen^16^, Karin I. Proper^16,19^, Maryam Kavousi^20^, W. M. Monique Verschuren^19,21^, Peter H. Bisschop^12^, Petra Juffer^22^, Richard Jaspers^23^, Sandra H. van Oostrom^19^, Sarah E. Siegelaar^12^, H. Susan J. Picavet^19^

^7^Leiden University Medical Center, Leiden University, Center for Bone Quality, Department of Endocrinology, Leiden, Amsterdam, The Netherlands

^8^Amsterdam University Medical Center, University of Amsterdam, Department of Laboratory Medicine, Endocrine Laboratory; Gastroenterology Endocrinology & Metabolism; Reproduction & Development, Amsterdam, The Netherlands

^9^Amsterdam University Medical Center, Vrije Universiteit Amsterdam, Department of Laboratory Medicine, Endocrine Laboratory; Gastroenterology Endocrinology & Metabolism; Reproduction & Development, Amsterdam, The Netherlands

^10^Academic Centre for Dentistry Amsterdam, University of Amsterdam and Vrij Universiteit, Department of Oral Cell Biology, Amsterdam, The Netherlands

^11^Hogeschool Utrecht, Utrecht, The Netherlands

^12^Amsterdam University Medical Center, University of Amsterdam, Department of Endocrinology, Amsterdam Gastroenterology Endocrinology Metabolism, Amsterdam Movement Sciences, Amsterdam, The Netherlands

^13^Athena Institute, Faculty of Science, Vrije Universiteit, Amsterdam, The Netherlands

^14^ACTA, faculty of Dentistry, University of Amsterdam, Amsterdam, The Netherlands

^15^Erasmus MC University Medical Center, Department of Internal Medicine, Rotterdam, The Netherlands

^16^Amsterdam UMC, University of Amsterdam, Department of Public and Occupational Health, Amsterdam Public Health, Amsterdam, The Netherlands

^17^Erasmus MC University Medical Center, Erasmus MC Cardiovascular Institute, Rotterdam, The Netherlands

^18^Avans University of Applied Sciences, Breda, The Netherlands

^19^National Institute for Public Health and the Environment, Centre for Prevention, Lifestyle and Health, Bilthoven, the Netherlands

^20^Erasmus MC University Medical Center, Department of Epidemiology, Rotterdam, The Netherlands

^21^Julius Center for Health Sciences and Primary Care, University Medical Center Utrecht, Utrecht University, Utrecht, the Netherlands

^22^Saxion University of Applied Sciences, Academt of Life Science, Engineering and Design, Research group Applied Nanotechnology, Enschede, The Netherlands

^23^Laboratory for Myology, Department of Human Movement Sciences, Faculty of Behavioural and Movement Sciences, Vrije Universiteit Amsterdam, Amsterdam Movement Sciences, The Netherlands

